# Effects of Savoring Meditation on Positive Emotions and Pain-Related Brain Function: A Mechanistic Randomized Controlled Trial in People With Rheumatoid Arthritis

**DOI:** 10.1101/2023.09.07.23294949

**Authors:** Patrick H. Finan, Carly Hunt, Michael L. Keaser, Katie Smith, Sheera Lerman, Clifton O. Bingham, Frederick Barrett, Eric L. Garland, Fadel Zeidan, David A. Seminowicz

## Abstract

Positive emotions are a promising target for intervention in chronic pain, but mixed findings across trials to date suggest that existing interventions may not be optimized to efficiently engage the target. The aim of the current mechanistic randomized controlled trial was to test the effects of a single skill positive emotion-enhancing intervention called Savoring Meditation on pain-related neural and behavioral targets in patients with rheumatoid arthritis (RA). Participants included 44 patients with a physician-confirmed diagnosis of RA (n=29 included in fMRI analyses), who were randomized to either Savoring Meditation or a Slow Breathing control. Both meditation interventions were brief (four 20-minute sessions). Self-report measures were collected pre- and post-intervention. An fMRI task was conducted at post-intervention, during which participants practiced the meditation technique on which they had been trained while exposed to non-painful and painful thermal stimuli. Relative to Slow Breathing, Savoring significantly reduced experimental pain intensity ratings relative to rest (p<.001), increased cerebral blood flow in the ventromedial prefrontal cortex (vmPFC) and increased connectivity between the vmPFC and caudate during noxious thermal stimulation (z=2.3 voxelwise, FDR cluster corrected p=0.05). Participants in the Savoring condition also reported significantly increased positive emotions (*p*s<.05) and reduced anhedonic symptoms (*p*<.01) from pre-to post-intervention. These findings suggest that that Savoring recruits reward-enhancing corticostriatal circuits in the face of pain, and future work should extend these findings to evaluate if these mechanisms of Savoring are associated with improved clinical pain outcomes in diverse patient populations.

## Introduction

Observational studies consistently demonstrate that positive emotions acutely inhibit pain and are associated with favorable long-term outcomes among patients with chronic pain[18; 46]. However, treatments that aim to augment positive emotional states as a means of regulating chronic pain have produced mixed results, with a recent meta-analysis identifying an overall small pooled effect size[46]. The largest randomized controlled trial (RCT) of a general positive psychological intervention for chronic pain to date (N=360 knee osteoarthritis patients) did not improve the primary outcomes of pain severity and physical function[37]. In contrast, mindfulness-based interventions have been theorized to promote adaptive pain self-management at least partially via increased positive emotions and the neural processes that give rise to them [13]. This theoretical premise is supported by a large literature base showing that mindfulness is a potent modulator of positive emotions [22; 31; 40]. A small number of studies show that a combination of mindfulness meditation, cognitive-behavioral tools, and hypnosis reduces clinical pain via increased positive emotions and neurophysiological response to rewards [25; 28; 48]. Although these results are promising, because the mindfulness-based interventions tested to date commonly include a variety of ‘active ingredients’, the primary driver of treatment effects is not clear. Collectively, these results suggest that positive emotions are a promising target for intervention in chronic pain, but existing interventions may not be optimized to efficiently engage the target.

The present mechanistic RCT was designed to test the mechanistic target engagement of a single skill positive emotion-enhancing intervention in patients with rheumatoid arthritis (RA). To that end, we developed a novel meditation technique called Savoring Meditation, during which patients were trained to generate positive emotional memories, enjoy a multisensory experience of the positive emotions that arise from those memories, and maintain attention toward that positive emotional state during periods of elevated pain. Our approach was derived in part from the mindful savoring skills taught in the mindfulness-based intervention Mindfulness-Oriented Recovery Enhancement (MORE) [24], as well as prior experimental work on positive autobiographical memories. Recall of positive autobiographical memories is associated with increased positive emotions among healthy participants[5; 57] and patients with mood disorders[2; 47]. Additionally, attentional engagement with positive autobiographical memories increases function within corticostriatal circuitry[44; 58; 60]a neural reward processing network that includes the nucleus accumbens (NAc) and ventral medial prefrontal cortex (vmPFC), among other brain regions that reliably respond to positive emotion inductions (for a review, see: [62]). To control for non-specific mechanisms supporting placebo effects, we employed a validated Slow Breathing Meditation control condition. We hypothesized that the active practice of Savoring, relative to rest, would increase activity within the NAc as a primary region of interest, and other corticostriatal brain regions discovered via whole brain analysis. We also hypothesized that, relative to Slow Breathing, RA patients trained in Savoring Meditation would report increased positive emotions from pre-to post-intervention. Clinical pain and anhedonia were explored as secondary self-report outcomes.

## Methods

### Study Overview

This was a randomized clinical trial (NCT03975595) that began as a 3-arm trial that included a Mindful Breathing intervention arm in addition to Savoring and Slow Breathing. Enrollment began in November, 2019. However, due to significant disruptions associated with the COVID-19 pandemic, which shuttered study operations beginning in March of 2020 and extended into October 2020, the following protocol changes were made:

1) the Mindful Breathing arm (n=6 were randomized to that arm) was dropped and the study proceeded as a 2-arm trial comparing Savoring to Slow Breathing.
2) All study visits and assessments were conducted remotely.
3) The intervention was modified from in-person to remotely delivered (via Zoom)
4) Post-questionnaires were required to be delivered remotely (via RedCap) instead of in person due to activity restrictions at the MRI facility requiring that only MRI procedures, and not questionnaires, be administered.

Prior to the COVID-19 disruption, 15 participants were enrolled in the trial. All pre-COVID-19 procedures were conducted in-person. Upon resuming the trial in October, 2020, only the final outcome assessment (fMRI) was conducted in-person, and the remaining procedures (screening, consenting, baseline assessment, and intervention sessions) were conducted remotely. All pre-COVID-19 participants in the Savoring and Slow Breathing groups were included in final analyses.

The study was not re-powered after dropping the third arm because the primary comparator of interest to Savoring was the Slow Breathing control, not the Mindful Breathing condition. Enrollment concluded in October, 2021.

### Participants

Participants included 44 randomized patients with a physician-confirmed diagnosis of RA. The full CONSORT diagram is presented in Figure 1, indicating that 50 participants were initially randomized, 6 of whom were included in the treatment arm that was subsequently dropped due to COVID-19. The CONSORT diagram also indicates that 15 Savoring participants and 14 Slow Breathing participants were included for primary fMRI outcome analysis, whereas all participants who provided baseline self-report data (21 Savoring participants and 23 Slow Breathing participants) were included in intent-to-treat analyses of self-report data.

**Figure 1.**
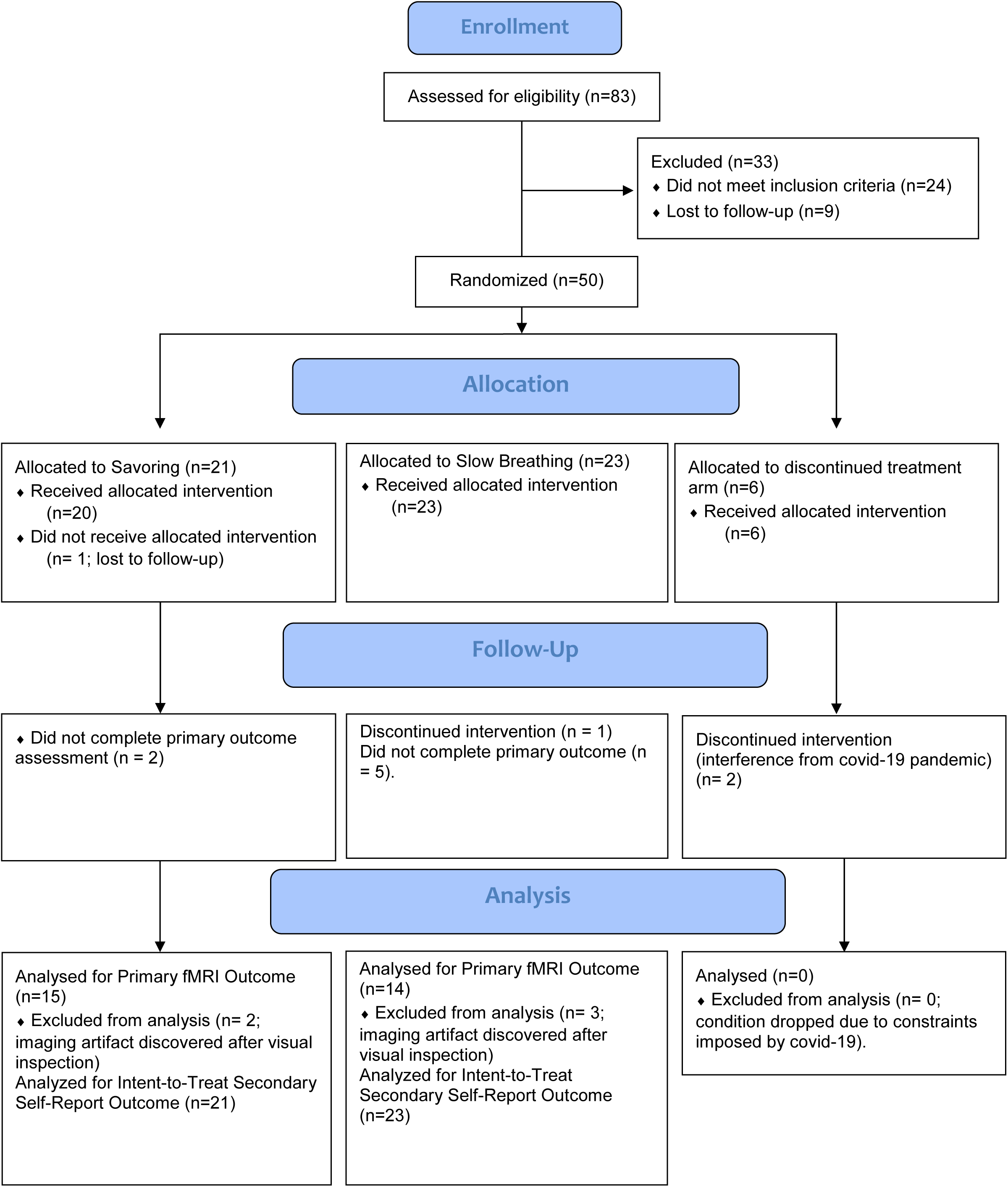
Consort Diagram

Patients were predominantly recruited from the Johns Hopkins Arthritis Center. Advertisements and fliers were also placed at community arthritis clinics and in social media forums. All participants provided informed consent, as required by the Johns Hopkins Institutional Review Board, which provided ethical approval for this study. The following inclusion/exclusion criteria were applied:

#### Inclusion Criteria

1) 30-70 years old; 2) have a physician-confirmed diagnosis of RA

#### Exclusion Criteria

1) unstable medical or psychiatric comorbidity within 3 months; 2) lifetime history of alcohol or substance use disorder; 3) current use of opioids (other than tramadol and/or codeine) for therapeutic or non-therapeutic purposes; 4) positive toxicology screen for recreational drugs; 5) pregnant or lactating women; 6) presence of any comorbid idiopathic (e.g., fibromyalgia) chronic pain condition; 7) prior meditation experience, with the exception of yoga; 8) cannot tolerate noxious thermal pain at any level above pain threshold; 9) contraindicated for MRI, assessed on an individual basis (e.g., metal devices, claustrophobia).

### Screening

Screening was conducted via questionnaires (administered through REDCap) and interview with the study coordinator. RA diagnosis and presence of exclusionary comorbidities were confirmed via medical record review and, if necessary, consultation with the study’s rheumatologist (CB).

### Interventions

#### Common Features

Interventions were conducted one-on-one between an interventionist and the participant. Both intervention conditions were structured to be brief and time-limited, with the goal of completing 4 separate sessions over a two week time period. Each session was scheduled for 30 minutes, with 5 minutes allotted at the beginning for introductory discussion, 20 minutes allotted for meditation, and 5 minutes allotted at the end for participants to ask questions. Both interventions had a similar structure, such that the first session included the greatest amount of interventionist guidance, with progressively less guidance in each subsequent session. Patients in both conditions were informed that the primary goal of the study was to evaluate what happens in the brain during meditation, and that after undergoing 4 sessions of meditation training, they would practice their newly acquired meditation skills in an MRI scanner during periods of rest and periods in which they were exposed to noxious thermal stimuli. As such, the meditation training was structured to progressively expose participants to elements of the scanning environment. After initially training participants in their assigned meditation practice in an upright posture during Sessions 1 and 2, participants were asked to meditate while lying supine for Sessions 3 and 4. Additionally, in Session 4, participants were exposed to scanner sounds (e.g., loud crackling noise with periodic chirping) that were recorded from a live fMRI scan. These additional features were introduced to ensure that participants were comfortable meditating in a scanning environment prior to undergoing their fMRI outcome assessment.

Because this was a mechanistic clinical trial and we sought to control intervention dose exposure as much as possible, participants were asked to refrain from home practice until after their participation was complete. Thus, meditation practice prior to the fMRI scan should have been equal across participants and limited to the four 20-minute sessions participants had with the interventionist.

Prior to the COVID-19 pandemic, intervention sessions were conducted in-person in a private room in our lab suite. Lab operations were shut down during the initial period of the pandemic, and the resumption of clinical research required implementing viral transmission risk mitigation. As such, we elected to conduct all further meditation training sessions over video conference (e.g., Zoom). The structure, timing, and content of the sessions did not change, and participants were assisted with technological details when needed.

#### Savoring

Savoring is a positive emotion-generative meditation practice through which participants cultivate a strong positive emotional state by drawing on a personally meaningful positive emotional memory. Individuals are first asked to identify a prior experience that brought about strong positive emotions when it occurred, and then are trained to attend to the memory of that experience and any sensory changes that occur when directing attention toward the memory. After utilizing the memory to evoke a positive emotional state, the meditation becomes focused on maintaining and increasing awareness of the multisensory experience of the positive emotional state in the present moment, including sensations of pleasure arising in the body. If practitioners experience mind wandering, they are encouraged to gently return attention to the present moment positive emotional experience and continue meditating by directing their awareness to that state. Aspects of the Savoring meditation training were adapted from savoring exercises included in Mindfulness-Oriented Recovery Enhancement[24], which teaches patients to savor positive emotional states but does not invoke positive emotional memories.

#### Slow Breathing

Slow Breathing is described to participants as a “breath-focused meditation.” The interventionist guides the participant to practice slow deep breathing while letting the body relax. However, the intervention does not explicitly train individuals in mindfulness skills, and at no time are participants instructed what to do with their thoughts or emotions during the meditation period. Mind-wandering is not acknowledged or discussed. Attention and awareness are not discussed. No explicit cognitive techniques are introduced. All other aspects (training setting, posture, facilitator, time providing instruction) of the Slow Breathing meditation intervention were matched to the Savoring meditation intervention. These intervention design choices were intended to distinguish this condition from mindfulness meditation. Slow Breathing was adapted from a similar active control condition in Zeidan et al.’s prior work[72].

### Randomization & Blinding

Participants were randomly assigned (50% probability) to Savoring or Slow Breathing. Participants, assessment technicians, and investigators were blind to treatment assignment, but the interventionists could not be blinded. To standardize expectations across conditions, Savoring and Slow Breathing were described to participants in general terms as “meditation-oriented” interventions. After completion of the study, participants were fully debriefed.

### Interventionists and Efforts to Promote Treatment Fidelity

The interventionists were two licensed clinical psychologists with prior training in meditation and pain psychology (CH and SL). They were trained and supervised in Savoring and Slow Breathing by the principal investigator (PHF). To promote fidelity to the interventions, both interventions were fully scripted and interventionists were trained to use the exact language included in each script for each session when leading patients in meditation. Review of a random subset of audio recordings confirmed that interventionists did, as expected, read the scripts as directed in the delivery of the interventions.

### Primary Outcome Measures: fMRI

#### fMRI Procedures

Scans were performed at the University of Maryland, Baltimore campus as soon as possible following the final intervention visit. At this visit (when feasible under COVID-19 protocols), participants underwent a QST familiarization session prior to the fMRI scan. During the familiarization session, participants were exposed to noxious thermal stimuli, delivered via Peltier thermode (Pathway, Medoc Inc.) to the calf, and trained on differentiating pain intensity ratings from pain unpleasantness ratings. The MRI scan session required approximately 75 minutes, but lasted up to 90 minutes in some participants who requested breaks due to joint discomfort. A standard anatomical T1-weighted scan (MPRAGE, voxels=1mm isotropic, TR=2300ms, TE=2.94ms, FA=9°, slices=192, scan time=5:30) was collected. fMRI scans used an arterial spin labeling sequence (ASL; Pseudocontinuous arterial spin labeling (PCASL)[53], single-shot EPI, whole brain, voxels=3×3, slice thickness=5mm, TR=4000ms, TE=13.0ms, FA=90°, slice gap=1mm, 20 slices, GRAPPA factor=2, scan time=4:08). ASL provides a quantifiable measurement of CBF (ml/100g tissue/min) that is accomplished by normalizing ASL 4D data by mean intensity values and designating each individual’s respective white matter value as a nuisance covariate in first-level analyses.[7; 49] Heart and respiratory rate data were collected during the scans for use in removing physiological noise from the fMRI data. There were four runs of rest and four runs in which participants are instructed to practice the skill learned in their respective intervention (Savoring, Slow Breathing). Thermal stimuli were delivered by a Pathway (Medoc Inc.) thermal stimulator to the calf. Thermal stimulus blocks were pseudorandomly interleaved in each set of the rest and “practice” runs so that runs alternated between warm non-painful and painful temperatures.

For the non-meditation runs, subjects were asked to not move, close their eyes, and stay awake. In meditation runs (Savoring and Slow Breathing), subjects were asked to “consciously relax the body, and invite yourself to let go of any tension you may be holding right now. Softening the muscles in the face, jaw and shoulders, and then relaxing all the way down the length of the body, relaxing the legs, feet and toes. Notice the points of contact between the body and the surface beneath you, and gently allow the eyes to close. Please begin your meditation practice now”. This instruction was given as a uniform instruction across conditions to help participants begin meditation from a relaxed state.

In all conditions, participants were asked to rate the pain intensity and unpleasantness associated with the thermal stimulus following each run. Following the fMRI procedures, participants completed questionnaires assessing levels of positive and negative emotions, and the extent to which they experienced mindfulness and/or savoring during the scan.

A noxious thermal stimulus task was chosen because prior work has shown that sensitivity to and descending modulation of noxious thermal stimuli discriminates patients rheumatoid arthritis from controls [1; 15; 64].

#### Target Brain Regions

The *a priori* region of interest (ROI) was the nucleus accumbens (NAc). ROIs for the NAc were created based on previous literature[4], derived from a voxel-wise contrast map of a heat pain rating task. NAC masks were created based on the center coordinate (16, 10, −8) specified in Balki et al [4] for R-NAC. An additional Left NAC mask was created (-16, 10, −8). Both left and right masks consisted of spheres with radius 5mm.

Although several brain regions were of interest, the NAc was chosen as the *a priori* ROI because our prior work showed that the NAc is modulated by both positive affective stimuli and noxious thermal stimuli [21]. However, we were also interested in other corticostriatal regions, including the vmPFC, but only followed up on them if they emerged as significant brain regions from whole brain analyses.

### Secondary Outcome Measures: Self-Report Questionnaires

#### Positive and Negative Affect

The Positive and Negative Affect Schedule-X (PANAS-X)[68] is a 60-item measure assessing perceived intensity of various specific emotions, which are aggregated to form positive and negative affect scales, and series of subscales (e.g., joviality).

#### Snaith-Hamilton Anhedonia and Pleasure Scale (SHAPS)

The SHAPS[56] is a 14-item measure assessing anhedonic symptoms (i.e., the inability to experience pleasure from naturally rewarding activities of daily life).

#### Treatment Credibility

Treatment credibility and satisfaction was assessed by modifying the Credibility and Expectancy Questionnaire[14] for use in a pain-focused intervention for RA patients. The following six items were assessed on an 11-point scale, where 0= Not at All and 10=Extremely: “How logical did this type of treatment seem to you?”; “How confident are you that this treatment would be successful in reducing rheumatoid arthritis-related pain?”; “How confident would you be in recommending this treatment to a friend who has rheumatoid arthritis?”; “How successful do you believe this treatment would be in helping you decrease the amount of interference that rheumatoid arthritis has in your life?”; “ How successful do you believe this treatment would be in helping you improve your mood?”; and “How successful do you believe this treatment would be in helping you improve your quality of life?” Scores were summed across items to achieve a total score of treatment credibility, with a maximum score of 60.

### Data Analysis

#### Statistical Approach for Patient-Reported Outcomes

Data were inspected for normality and outliers (+/- 3 *SD* around the mean). One outlier in SHAPS (i.e., anhedonia) scores was detected and removed. The effect of group assignment (Savoring vs. Slow Breathing) on affective outcomes was tested using intent-to-treat linear mixed-effects models allowing for random intercepts in accordance with recommended procedures in longitudinal analysis[55]. We first examined the effect of time (i.e., baseline to post-treatment) within Savoring and Slow Breathing groups separately. We then tested a group (Savoring vs Slow Breathing) by time (baseline vs post-treatment) interaction term for each outcome. Age, race and clinical pain severity are associated with individual differences in affective function and pain sensitivity in response to noxious stimuli [41; 50; 59; 69; 71]. Thus, we evaluated baseline correlations between these variables and the patient reported outcomes to determine covariate selection. Significant associations emerged between baseline pain and anhedonia (r = .39, p < .05), black race (vs. all others) and anhedonia (r = .37, p < .05), and black race (vs. all others) and joviality (r = .42, p < .05), and these variables were adjusted accordingly in regression models. <u>Thermal stimulus intensity was not significantly associated with pain ratings in models of experimental pain outcomes (p’s≥.06) and did not the magnitude of primary effects, and so was not included as a covariate in final models.</u> Models were specified using the lme4 and lmerTest packages [6; 43] in R using restricted maximum likelihood estimation. Random intercepts and slopes were both initially modeled, but convergence could not be achieved in models including random slopes, so final models presented reflect random intercepts only. Significance values were computed using Satterthwaite’s method[43]. We probed significant interaction effects by visualizing simple slopes. Within-group effect sizes (Cohen’s *d*’s) were computed by dividing the mean difference between pre and post-scores by the averaged standard deviation in accordance with recommended approaches[11].

Because 9 participants who enrolled prior to the COVID-19 pandemic completed their outcome assessments in-person and the remaining 35 participants completed their outcome assessments remotely due to in-person restrictions, we conducted sensitivity analyses with a dichotomous covariate indicating whether the participant completed assessments prior to or after the shift to remote assessment. The nature and significance of effects of these models was similar across all self-reported outcomes, so final models presented herein do not include the dichotomous covariate.

#### fMRI Pre-Processing

Imaging analysis was conducted using Functional Magnetic Resonance Imaging of the Brain (FMRIB) Software Library (Center for FMRIB, University of Oxford, Oxford, UK) and ASL data processing toolbox scripts provided by Chris Rorden (https://crnl.readthedocs.io/asl/index.html). Five subjects were excluded from analysis due to detected image artifact or excessive motion. A total of 29 subjects (15 Savoring, 14 Slow Breathing) were included in the final imaging analysis. Subjects underwent 8 functional (ASL) runs during the MRI challenge session. The 8 ASL images that resulted from the runs represented combinations of two experimental conditions: Pain (nonpainful warm stimulus vs. noxious heat) and Practice (no meditation vs. meditation). These combinations occurred in the following order: 1. warm + no meditation; 2. noxious + no meditation; 3. warm + no meditation; 4. noxious + no meditation; 5. warm + meditation; 6. noxious + meditation; 7. warm + meditation; 8. noxious + meditation. “No Meditation” conditions were included before “Meditation” conditions to avoid carryover effects of meditation. Notably, there was no non-warm baseline, so the Warm condition served as the primary comparator to the Pain condition. ASL images were motion corrected using the ASL MRI MoCo Method[66]. An additional motion correction and signal outlier filtering step was included[63] to account for excessive motion and corruptive signal fluctuations. ASL images were converted into absolute CBF units according to the general kinetic model. The 8 CBF images were concatenated, intensity normalized, and then smoothed with a 9mm full-width half maximum (FWHM) Gaussian kernel. The images were spatially normalized to the MNI (Montreal Neurologic Institute) standard space using a 12-parameter affine transformation and then a nonlinear transformation. Additionally, a global white matter value was calculated for each CBF image by extracting average white matter values from FSL’s MNI152 white matter mask. The global white matter value was included in statistical models as a nuisance regressor. **<u>Statistical Approach for fMRI:</u>** As a pilot mechanistic trial, the study was powered to detect a large effect (d=.9) with at least 16 participants per group. A group one-sample t-test (n = 29) was used to determine whole-brain, pain-related regions. The two Warm+No Meditation scans were subtracted from the two Noxious+No Meditation scans and then averaged. This average residual value per subject was used as the dependent measure included in the one-sample t-test.

To determine the effects of Pain and Practice per subject, a 2 (Pain) x 2 (Practice) repeated measures design was used. First, voxelwise parameter estimate maps for the main effects for Pain, Practice and the Pain x Practice interaction were obtained for the *a priori* ROI, NAc. Next, whole-brain voxelwise parameter estimate maps were also obtained for the same contrasts. This was done separately for the Savoring and Slow Breathing groups. Third-level analyses compared the Savoring and Slow Breathing group main effects and interaction.

Multiple regression analyses investigated the relationship between pain intensity/unpleasantness ratings and CBF during the Noxious condition. The 1^st^ regressor consisted of the mean difference between the 2 Noxious+no Meditation scans vs Noxious+Meditation scans. The 2^nd^ regressor included the percent change in subjects’ pain intensity ratings from the Noxious+no Meditation to the Noxious+Meditation scans. The 3^rd^ regressor included the percent change in the subjects’ pain unpleasantness ratings from the Noxious+no Meditation to the Noxious+Meditation scans. All 3 regressors were orthogonalized to each other.

Because this was a proof-of-concept pilot mechanistic trial with a limited sample size, we set voxelwise threshold to 2.3, cluster corrected at a p=0.05 threshold [70], in line with previous meditation-based imaging studies of similar size[72].

##### Exploratory Functional Connectivity of VMPFC

Functional Connectivity analysis was conducted using The CONN Functional Connectivity Toolbox (version 21.a, https://www.nitrc.org/ projects/conn/) and ASL Data Processing Toolbox [67]. Preprocessing and analysis steps used were in accordance with methods described in previous literature[39]. ASL images were motion corrected, co-registered, segmented into CSF, WM, and GM images, and then spatially normalized to MNI space, adapting the ASL Toolbox preprocessing scripts provided by Chris Rorden (https://crnl.readthedocs.io/asl/index.html). Following methods described in Ibinson et al. [39], reprocessed images were then smoothed with 8 mm FWHM and denoised for WM, CSF, and effect for rest within Conn Toolbox. A 3mm radius sphere centered at the left ventromedial PFC (VMPFC; 14, 48, -4) coordinate was chosen from the result of the previously described 2^nd^-evel whole brain analysis for the Group by Practice by Pain triple interaction activation maps (described in Results section *Interaction of Meditation Practice and Pain on Whole Brain CBF*). The VMPFC seed-to-voxel maps for both the Savoring and Slow Breathing groups were produced with a cluster-forming threshold of p = 0.001 and FDR cluster correction p = 0.05.

## Results

### Participants

Table 1 provides demographic information for all study participants. At baseline, Savoring and Slow Breathing groups were undifferentiated on age (B = .89, se = 3.44, p =.80). Omnibus chi square tests suggested no significant between-groups differences on demographic characteristics as presented in Table 1, including race (chi square = .44, df = 3, p = .93), Hispanic ethnicity (chi square = .46, df = 1, p = .50), income (chi square = 2.27, df = 3, p = .52), sex (chi square = .27, df = 1, p = .61), and education (chi square = 4.79, df = 3, p = .19). Across the full sample, baseline pain severity (M=3.19, SD=2.29) and pain interference (M=2.31, SD=2.54) were generally low. The majority of participants were prescribed disease-modifying anti-rheumatic drugs (DMARDs), and were required to be on stable doses (at least 4 weeks) prior to enrollment. Across the full sample, 66% (n=33) of patients were prescribed non-biologic DMARDs and 62% (n=31) were prescribed biologic DMARDs. Broken down by treatment group, 57.14% (n=12) of patients in the Savoring group were prescribed non-biologic DMARDs and 61.90% (n=13) were prescribed biologic DMARDs. In the Slow Breathing group, 78.26% (n=18) of patients were prescribed non-biologic DMARDs and 60.87% (n=14) were prescribed biologic DMARDs. All patients remained on the same DMARD medications at the same dose throughout the trial.

**Table 1.** Participant demographics by group.

|  | Savoring<br>Meditation | Slow<br>Breathing |
| --- | --- | --- |
| Total $n$ | 21 | 23 |
| Age, $M$ ($SD$) | 50.7 (12.0) | 49.8 (10.9) |
| Female, $n$ (%) | 20 (95) | 21 (91) |
| Education, $n$ (%) | | |
| High School | 6 (29) | 6 (26) |
| Undergraduate<br>Degree | 5 (24) | 9 (39) |
| Graduate degree | 9 (43) | 4 (17) |
| M.D. | 1 (5) | 4 (17) |
| Race, $n$ (%) | | |
| Asian | 1 (5) | 2 (9) |
| Black or African<br>American | 5 (24) | 5 (22) |
| White or European<br>American | 13 (62) | 13 (57) |
| Other | 2 (10) | 3 (13) |
| Hispanic or<br>Latino/a, $n$ (%) | 2 (10) | 1 (4) |
| Income, $n$ (%) | | |
| < 20,000 | 3 (14) | 1 (4) |
| 20-60,000 | 3 (14) | 6 (26) |
| 60-100,000 | 3 (14) | 2 (9) |
| 100,000 or more | 12 (57) | 14 (61) |

No participants were taking opioid analgesics, per protocol. Across the entire sample, 44% of patients were reported non-steroidal anti-inflammatories (NSAIDs), with similar rates in the Savoring (42.86%; n=9) and Slow Breathing (43.48%; n=10) groups.

### Intervention Feasibility, Credibility, and Adverse Events

All patients who initiated the interventions (both Savoring and Slow Breathing) completed 100% of intervention sessions. The goal was for patients to complete the 4 intervention sessions over the course of 2 weeks. The average time to complete all intervention sessions was 22.08 days (SD = 9.78).

Participants responded to the item, “How much do you expect meditation to reduce your pain?” (0 = not at all; 100 = completely) at baseline. Savoring (M = 55.0, SD = 26.0) and Slow Breathing groups (M = 54.8, SD = 25.4) were undifferentiated on baseline meditation expectations, as indicated by a linear regression model regressing group assignment on meditation expectations (B = .21, se = 7.75, p = .98).

Patients found both Savoring and Slow Breathing to be credible interventions. Patients in both groups reported a high level of confidence in each intervention’s potential to treat RA pain (Savoring: M = 7.05, SD = 1.90; Slow Breathing: M = 7.27, SD = 2.14; *p* = .73). Overall treatment credibility was high, and equivalent between groups (Savoring: M = 46.50, SD = 7.07; Slow Breathing: M = 46.10, SD = 10.10; *p* = .89).

There were no adverse events reported with either Savoring or Slow Breathing.

### Acute Effects of Active Savoring Meditation on Experimental Pain Sensitivity

#### Pre-Scan Thermal Stimuli Ratings and Limits

Prior to entering the fMRI scanner, participants were exposed to varying levels of thermal stimuli to determine individually tailored noxious temperatures for use during the scan. The average pre-scan temperature eliciting a pain intensity rating within the pre-specified window for inclusion (between 50 and 70/100) was 45.07°C (SD = 1.60).

#### Application of Thermal Stimuli During Rest versus Meditation

In the Savoring group, noxious thermal stimuli applied during rest resulted in average pain intensity ratings of 44.93 (SD = 16.74) and pain unpleasantness ratings of 37.72 (SD = 16.39). Active Savoring during the presentation of noxious thermal stimuli significantly reduced pain intensity by 30.46% (b = 13.70, se = 3.25, p < .001; M = 31.24.88; SD = 13.89) and pain unpleasantness by 40.67% (b = 15.34, se = 4.05, p= .002 M = 22.38; SD = 13.34), relative to rest.

Among participants in the Slow Breathing group, noxious thermal stimuli applied during rest were associated with average pain intensity ratings of 53.31 (SD = 11.08) and pain unpleasantness ratings of 45.51 (SD = 19.89). Active Slow Breathing during the presentation of noxious thermal stimuli significantly reduced pain intensity by 19.86% (b = 10.57, se = 4.27, p = .02; M = 42.74; SD = 21.46) and pain unpleasantness by 23.56% (b = 10.72, se = 4.83, p = .04; M = 34.79; SD = 25.94), relative to rest.

There was a marginally significant between-person main effect of Group on pain intensity (b = - 9.94, se = 4.89, p = .05), such that pain ratings in response to noxious heat applied during meditation were lower overall among patients in the Savoring group compared to the Slow Breathing group. However, the Group (Savoring vs. Slow Breathing) X Condition (Practice vs. Rest) interaction was not significant (b = 3.12, se = 5.36, p = .57).

Together, these findings indicate that both Savoring and Slow Breathing practices were acutely analgesic to noxious thermal stimuli, but that pain sensitivity overall was lower during Savoring compared to Slow Breathing.

### Manipulation Check for Pain-Related Brain Activation

#### Pain-related cerebral activation

We first evaluated the effects of noxious stimuli to determine pain-related brain activity maps across groups with a one-sample t-test. The Pain condition (noxious versus warm) was associated with increased CBF in right anterior insula (aIns), bilateral ventrolateral prefrontal cortex (VLPFC), right dorsolateral PFC (DLPFC), right superior parietal lobule (SPL), and decreased CBF in bilateral visual cortex, bilateral precuneus, and left superior temporal gyrus (because the extent of activations at z=2.3 was very large and encompassed multiple anatomic locations, interpretation of the findings was better facilitated at z=3.1, cluster corrected p=0.05 for this analysis; Figure 2 demonstrates effects at both z=2.3 and z=3.1 to show the extent of activation). These effects were less pronounced in the individual groups: increased CBF was seen in the right aIns in both groups, but left aIns, right SPL, and left DLPFC increases were seen only in the Savoring group, and right DLPFC increase was only seen in the Slow Breathing group (z=2.3 voxelwise, cluster corrected p=0.05; Table 2 “Noxious Heat Effect-Savoring Group; Noxious Heat Effect-Slow Breathing Group”).

**Figure 2.**
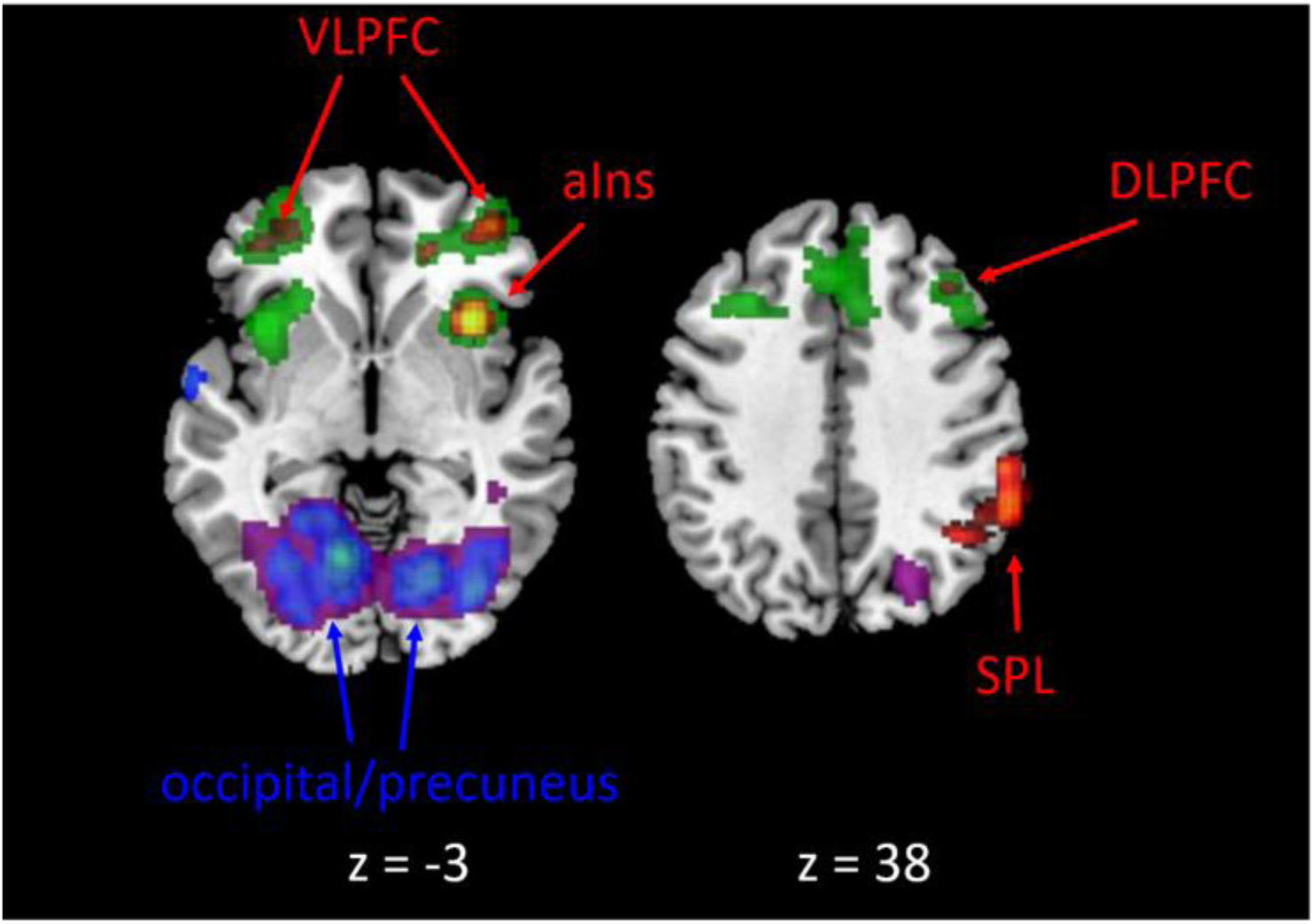
CBF changes associated with pain, across groups while not practicing meditation. Underlay (increases in green, decreases in purple) shown at z=2.3, cluster corrected p=0.05. Overlay (increases in red, decreases in blue) shown at z=3.1, cluster corrected p=0.05. z refers to MNI coordinates. Left side of brain is left. aIns, anterior insula cortex; VLPFC, ventrolateral prefrontal cortex; DLPFC dorsolateral prefrontal cortex; SPL, superior parietal lobule.

**Table 2.** fMRI statistics and coordinates.

| ROI | Volume (mm<sup>3</sup>) | Peak Z | MNI Coordinates (xyz) |
| --- | --- | --- | --- |
| **Noxious Heat Effect-Full Sample** |  |  |  |
| **Increased CBF** |  |  |  |
| Right anterior insula cortex | 3496 | 5.0499 | 38 16 2 |
| Left ventrolateral prefrontal cortex | 2288 | 3.9559 | -36 42 0 |
| Right superior parietal lobule | 1744 | 4.214 | 52 -40 58 |
| Right ventrolateral prefrontal cortex | 776 | 3.9758 | 32 46 -10 |
| Right dorsolateral prefrontal cortex | 800 | 4.5272 | 40 30 34 |
| Right putamen | 160 | 3.5726 | 26 20 0 |
| **Decreased CBF** |  |  |  |
| Left occipital cortex | 1872 | 5.4306 | -8 -64 -6 |
| Right occipital cortex | 1720 | 5.2254 | 34 -78 -6 |
| Left fusiform gyrus | 1168 | 4.0779 | -32 -68 -4 |
| Right fusiform gyrus | 224 | 3.1826 | 26 -58 -16 |
| Left precuneus | 104 | 3.2871 | -12 -52 20 |
| Right precuneus | 160 | 3.1619 | 2 -60 20 |
| Left superior temporal gyrus | 104 | 3.1632 | -66 -8 0 |
| **Noxious Heat Effect-Savoring Group** |  |  |  |
| **Increased CBF** |  |  |  |
| Right anterior insula cortex | 1888 | 5.192 | 36 22 4 |
| Left anterior insula cortex | 576 | 3.9168 | -26 16 -14 |
| Left dorsolateral prefrontal cortex | 2400 | 6.8327 | -36 20 38 |
| Right superior parietal lobule | 1264 | 3.9859 | 42 -60 50 |
| **Decreased CBF** |  |  |  |
| Left occipital cortex | 1472 | 5.0788 | -18 -62 -8 |
| Left Fusiform Gyrus | 680 | 4.3894 | -28 -62 -12 |
| Right occipital cortex | 432 | 3.7621 | 28 -56 -4 |
| Left Precuneus | 32 | 3.028 | -2 -44 14 |
| Right Precuneus | 80 | 3.0739 | 2 -64 14 |
| **Noxious Heat Effect-Non-Mindful Breathing Group** |  |  |  |
| **Increased CBF** |  |  |  |
| Right anterior insula cortex | 2224 | 8.5548 | 28 28 8 |
| Right dorsolateral prefrontal cortex | 320 | 6.4209 | 40 24 26 |
| Right Putamen | 128 | 4.8736 | 28 12 6 |
| **Decreased CBF** |  |  |  |
| Left occipital cortex | 2016 | 6.6012 | -2 -70 14 |
| Right occipital cortex | 960 | 6.802 | 6 -78 22 |
| Left Precuneus | 688 | 5.2564 | -14 -46 4 |
| Right Inferior Temporal Gyrus | 272 | 4.0948 | 54 -66 6 |
| **Practice Effect-Savoring Group** |  |  |  |
| **Decreased CBF** |  |  |  |
| Left superior temporal/S1/M1 | 14768 | 9.3907 | -62 -6 24 |
| Right superior temporal/S1/M1 | 7336 | 6.6701 | 60 -14 4 |
| **Practice Effect-Non-Mindful Breathing Group** |  |  |  |
| **Decreased CBF** |  |  |  |
| Left occipital cortex | 7616 | 9.9665 | -46 -80 -8 |
| **Pain X Practice Effect-Savoring Group** |  |  |  |
| **Increased CBF** |  |  |  |
| midbrain/PAG, brainstem, pons, occipital | 13952 | 7.3351 | -14 -68 14 |
| **Pain Intensity Effect-Savoring Group** |  |  |  |
| **Increased CBF** |  |  |  |
| Left frontal operculum | 6848 | 6.2796 | -56 16 14 |
| Left posterior insula | 304 | 2.7344 | -30 -2 14 |
| **Decreased CBF** |  |  |  |
| Left occipital cortex | 20320 | 7.4523 | -22 -82 24 |
| Left middle temporal gyrus | 10200 | 6.584 | -56 -24 -12 |
| Right precuneus | 376 | 3.8235 | 14 -50 26 |
| Right occipital cortex | 40 | 3.4142 | 20 -78 10 |
| **Group x Pain x Practice Effect** |  |  |  |
| **Increased CBF in SAV vs NMB** |  |  |  |
| Left ventromedial prefrontal cortex/<br>pregenual anterior cingulate cortex | 11720 | 5.8902 | -14 48 -4 |
| **Seed-to-voxel connectivity of VM PFC** |  |  |  |
| Left caudate | 1016 | 7.2225 | -14 10 4 |

#### Relationship between pain intensity/unpleasantness ratings and CBF

During both the Meditation and No Meditation conditions, the Savoring group’s pain intensity ratings were associated with increased CBF in left posterior insula and frontal operculum, and decreased CBF in visual cortices (z=2.3 voxelwise, cluster corrected p=0.05; Table 2 “Pain Intensity Effect-Savoring Group”).

### Effects of Savoring Meditation on Brain Function

#### Primary Analyses of Meditation Practice Effects on NAc

In the Savoring group, the main effect of Meditation (vs. No Meditation) was not significant for lNAc (b = -242.59, se = 243.59, p = .34) or rNAc (b = -19.50, se = 272.69, p = 0.94) activation during noxious stimulation. Similarly, the main effect of Slow Breathing (vs. No Meditation) was not significant for lNAc (b = -216.05, se = 453.38, p = 0.64) or rNAc (b = 631.51, se = 483.71, p = 0.22) activation during noxious stimulation. The group X practice interaction was not significant for lNAc (b = 4.23, se = 510.90, p = 0.99) or rNAc activation (b = -632.33, se = 550.18, p = 0.26) during noxious stimulation. The group X practice X pain interaction was not significant for lNAc (b = -295.24, se = 615.25, p = 0.63) or rNAc (b = 660.26, se = 792.02, p = 0.41) activation.

#### Secondary Analyses of Meditation Practice Effects on Whole Brain CBF

A main effect of Savoring Meditation (vs. No Meditation) was associated with decreased CBF in bilateral superior temporal cortex and right S1/primary motor cortex (M1) (Table 2 “Practice Effect-Savoring Group”). Slow Breathing was associated with decreased CBF in left visual cortex (Table 2 “Practice Effect-Slow Breathing Group”). A Group (Savoring vs. Slow Breathing) by Practice (Meditation vs. No Meditation) interaction revealed that the effect of practice on CBF in right superior temporal cortex/S1/M1 was significantly greater in Savoring compared to Slow Breathing (z=2.3 voxelwise, cluster corrected p=0.05).

#### Interaction of Meditation Practice and Pain on Whole Brain CBF

For Savoring, Pain X Practice effects were seen as increased CBF in midbrain (including the PAG), brainstem, pons, and occipital cortex (z=2.3 voxelwise, cluster corrected p=0.05; Table 2 “ Pain X Practice Effect-Savoring Group”). For Slow Breathing, no significant Pain X Practice effects emerged.

A Group X Practice X Pain interaction showed that participants in the Savoring group had increased left vmPFC during Noxious stimuli compared to Slow Breathing (z=2.3 voxelwise, cluster corrected p=0.05; Figure 3; Table 2 “Group X Pain X Practice Effect”). The vmPFC cluster spread posteriorly to include pregenual ACC (pACC), and extended into caudate and OFC. Although the current study did not have the statistical power to resolve these areas as unique activations, the findings are highly suggestive of involvement of this wider reward/value appraisal circuitry.

**Figure 3.**
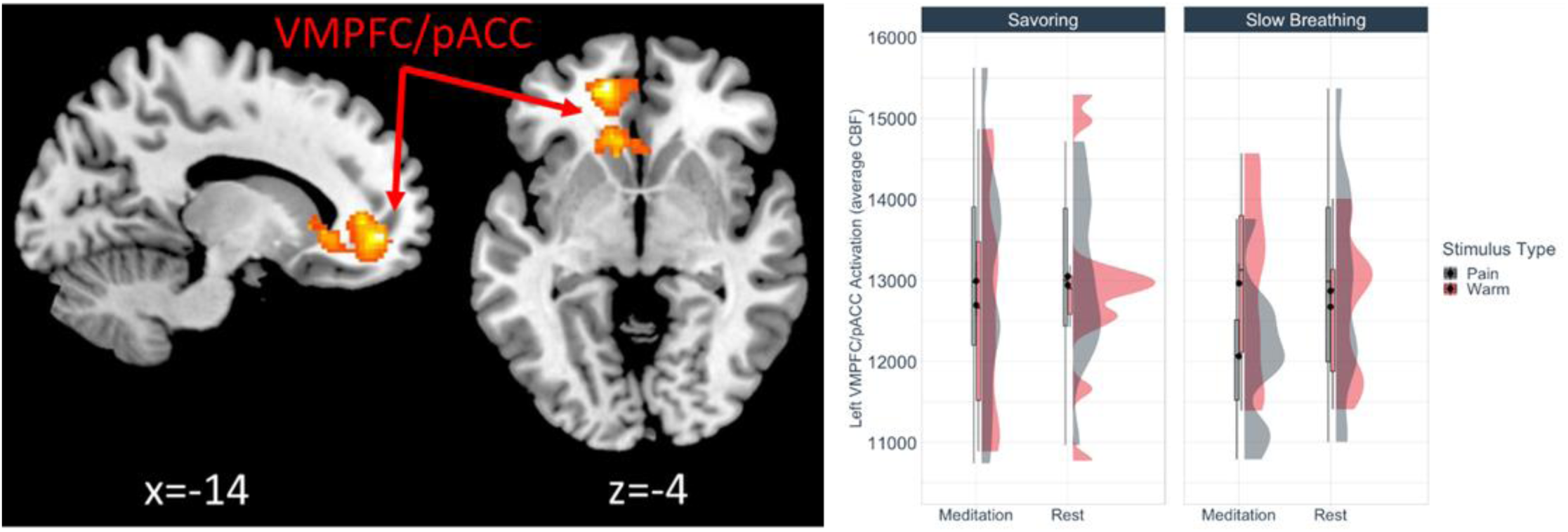
Left VMPFC/pACC was activated by savoring during pain. Based on a triple interaction (Group x Practice x Pain). z=2.3, p=0.05 cluster corrected. VMPFC/pACC, ventromedial prefrontal cortex/pregenual anterior cingulate cortex. The plot to the right shows the mean and median CBF values, along with distributions, as a function of Group, Practice, and Stimulus type.

#### Exploratory Connectivity of vmPFC

As a post-hoc analysis, we further investigated the connectivity of vmPFC as it fit the hypothesis that Savoring would engage corticostriatal circuitry beyond the NAc. We estimated time-series based whole brain connectivity using vmPFC as the seed region (using the peak voxel and a 3mm sphere). We found increased connectivity between left vmPFC and left caudate for Savoring during pain [z=2.3 voxelwise, FDR cluster corrected p=0.05; Figure 4 (z=3.1 overlaid); Table 2 Seed-to-voxel connectivity of vmPFC). This connectivity was not present for Slow Breathing. This caudate cluster survived direct comparison between Savoring and Slow Breathing at uncorrected threshold of p=0.001. These findings suggest that during noxious stimulation, practicing Savoring engages a descending pain modulatory response via vmPFC that is not seen during Slow Breathing.

**Figure 4.**
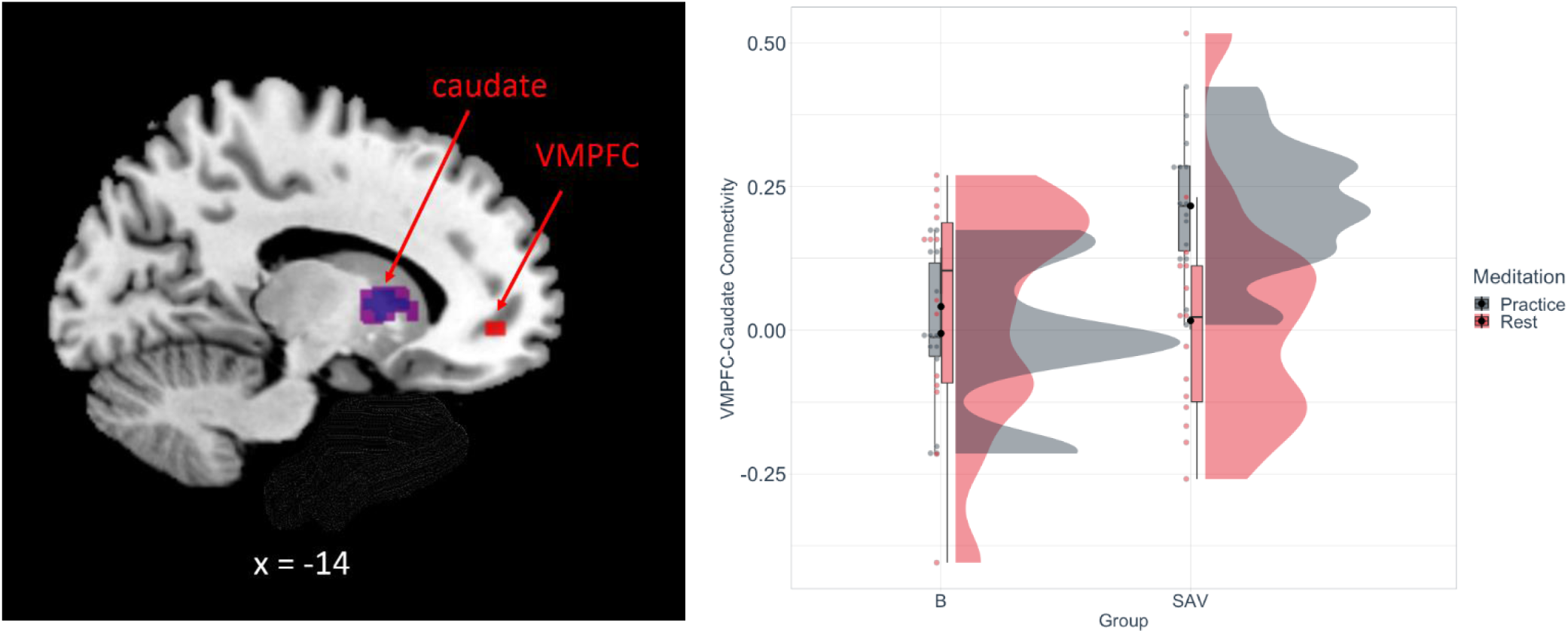
Seed-to-voxel analysis of the vmPFC (seed shown in red) revealed connectivity with the left caudate nucleus during Savoring while in pain. The plot to the right shows the connectivity strength between groups with and without meditation. z=2.3, p=0.05 cluster corrected (purple), z=3.1, p=0.05 cluster corrected (blue). vmPFC, ventromedial prefrontal cortex.

### Effects of Savoring Meditation Training on Pre-to-Post Measures of Affect and Clinical Pain

Although the trial was designed primarily to investigate the effects of active Savoring Meditation on brain function during an evoked pain task, baseline and post-treatment measures of positive affect, negative affect, anhedonia, and clinical pain were obtained for secondary analyses. As a greater number of participants completed these measures than provided usable imaging data, we included the full sample (N=44) for these analyses. Table 3 provides pre- and post-intervention means and standard deviations for key affective variables.

**Table 3.** Pre- and Post-Intervention Descriptive Statistics for Affective Measures.

|  | Pre-Treatment |  | Post-Treatment |  | p | Cohen's *d* |
| --- | --- | --- | --- | --- | --- | --- |
|  | *M* | *SD* | *M* | *SD* |  |  |
| Savoring (N=21) |  |  |  |  |  |  |
| Generalized PA | 32.62 | 8.35 | 37.33 | 7.63 | .06 | .59 |
| Joviality | 23.95 | 7.36 | 29.50 | 7.81 | .007 | .73 |
| Serenity | 9.00 | 2.17 | 10.39 | 2.59 | .04 | .58 |
| Generalized NA | 14.71 | 4.06 | 14.72 | 5.15 | .47 | .002 |
| Anhedonia | 19.76 | 5.45 | 16.22 | 3.06 | .006 | .83 |
| Slow Breathing<br>(N=23) |  |  |  |  |  |  |
| Generalized PA | 31.61 | 7.67 | 30.83 | 6.56 | .67 | .11 |
| Joviality | 23.17 | 5.97 | 22.89 | 6.60 | .65 | .05 |
| Serenity | 9.83 | 2.04 | 9.67 | 1.88 | .56 | .08 |
| Generalized NA | 14.65 | 4.73 | 15.11 | 4.10 | .98 | .10 |
| Anhedonia | 20.39 | 9.17 | 19.17 | 4.74 | .90 | .18 |

Among patients in the Savoring group, there was a significant effect of Time on the specific positive emotion subscales joviality (b = 5.11, se = 1.67, p = .007) and serenity (b = 1.25, se = 0.56, p = .037), with positive emotions in each domain increasing from pre-to post-intervention. General positive affect showed similar, but non-significant improvement (b = 4.33, se = 2.14, p = .057) following Savoring. Patients in Slow Breathing did not report significant changes in general positive affect (b = -0.59, se = 1.36, p = .67), joviality (b = .47, se = 1.03, p = .65), or serenity (b = -.20, se = .35, p = .56). There were significant Group X Time interactions for joviality (b = -4.89, se = 1.96, p = .02) and serenity (b = -1.43, se = .65, p = .04), indicating that the change from pre-to post-intervention was significantly greater in the Savoring group compared to Slow Breathing. The Group X Time interaction for general positive affect was not statistically significant (b = -4.80, se = 2.52, p = .065). Figure 5 (panels A, B and C) displays the results of these models.

**Figure 5.**
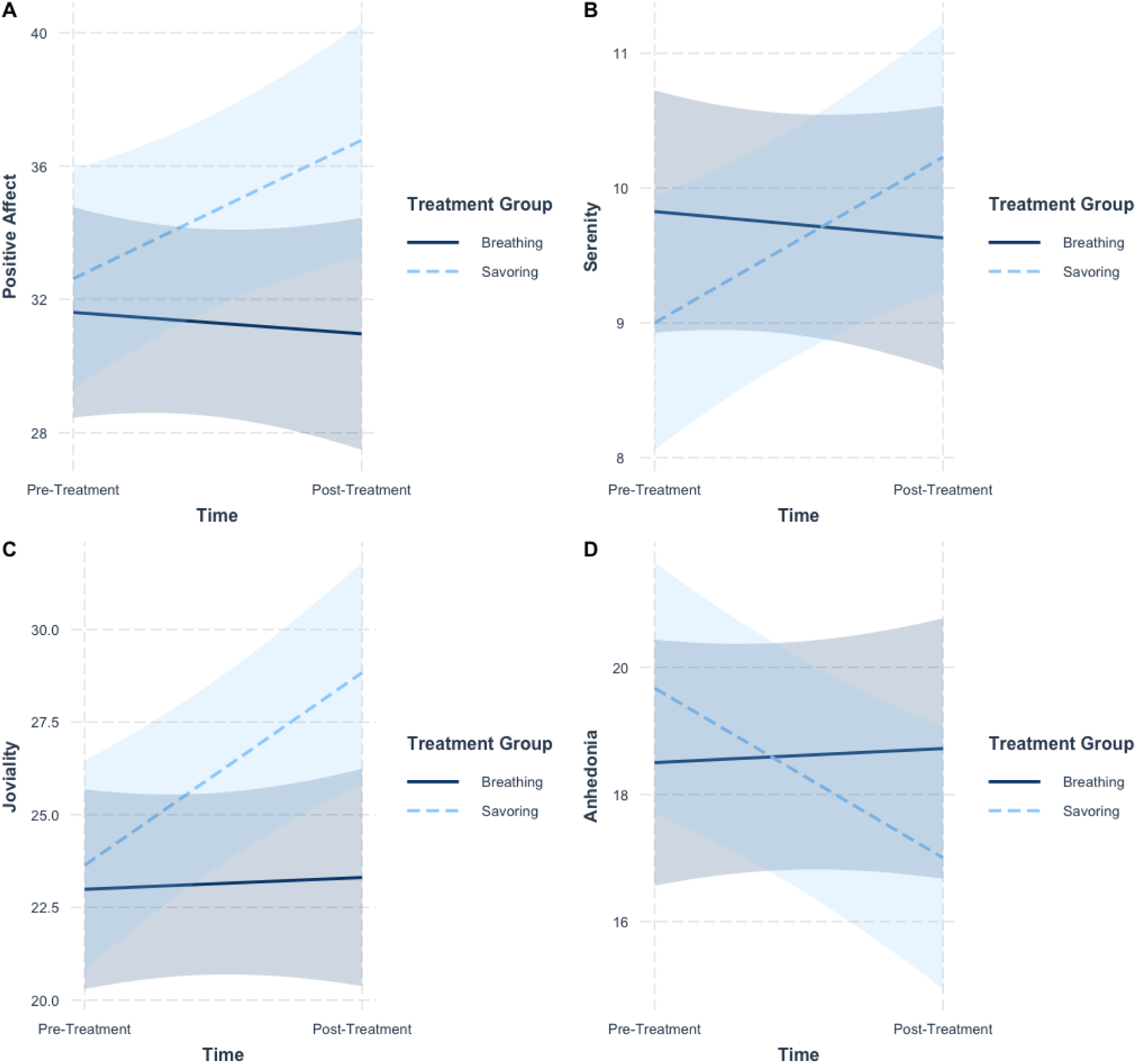
Pre-to post-treatment group differences between Savoring (SAV) and Slow Breathing on general Positive Affect (Panel A), Serenity (Panel B); Joviality (Panel C); and Anhedonia (Panel D).

Neither Savoring (b = 0.60, se = 0.81, p = .47) nor Slow Breathing (b = -0.03, se = .94, p = .98) altered negative affect from pre-to post-intervention, and the Group X Time interaction was not significant (b = -0.63, se = 1.25, p = .62).

Patients in Savoring reported significant pre-to post-intervention decreases in anhedonia symptoms (b = -2.59, se = 0.83, p = .006). Anhedonia did not significantly change in the Slow Breathing group (b = .11, se = .90, p = .90). The Group X Time interaction was significant (b = 2.89, se = 1.23, p = .03), indicating that the change in anhedonia symptoms from pre-to post-intervention was significantly greater in the Savoring group than the Slow Breathing group. Figure 5 (panel D) displays this finding.

Linear mixed models revealed that clinical pain severity did not significantly change over time in either the Savoring Group (B = -0.15, se = .25, t(16.49) = -0.63, p = .54) or the Slow Breathing Group (B = -0.08, se = .35, t(17.6) = -0.24, p = .82). The Group X Time interaction was also not significant (B = .15, se = .43, t(33.56) = .33, p = .74).

## Discussion

The primary findings of the present study of patients with RA are that Savoring Meditation is acutely analgesic, modulates CBF within corticostriatal circuitry during noxious stimulation, and leads to increases in positive emotions and decreases in anhedonic symptoms without altering negative emotions. On those target-specific secondary measures, Savoring outperformed a strong, active Slow Breathing control intervention, despite the fact that patients found both conditions highly credible as potential clinical interventions for RA.

Savoring was developed as a time- and content-limited intervention to augment positive emotional function and promote positive emotional pain inhibition in patients with chronic pain. This pilot RCT was designed to assess these mechanisms through both fMRI of brain response to active Savoring, and changes in patient reported outcomes following 4 brief Savoring training sessions. At this initial stage of iterative testing, the observations that Savoring increases positive emotions, is acutely analgesic, and engages brain regions that are implicated in cognitive-affective regulation of pain support further evaluation of the efficacy of Savoring as a brief pain self-management intervention. Notably, negative affect was not altered by Savoring. This may have been due to strong target specificity focal to positive emotions (see also: [22]), or alternatively may have been due to the relatively low levels of negative affect endorsed by the study sample creating a floor effect. Prior work has shown that positive and negative affect do not always move in lockstep in patients with chronic pain, with correlations in these studies typically falling in the r=.2 to r=.3 range [18]. Future studies will need to investigate the effects of Savoring on affective outcomes in patients with more severe affective disturbances to evaluate if patients with higher baseline levels of negative affect experience reductions concomitant with increases in positive affect.

The effect sizes of evidence-based pain self-management interventions are generally small[16; 65], and may be non-specific [8; 9], suggesting that they may need to be tailored to individual phenotypes to maximize benefit[12]. Interventions such as Savoring that explicitly target positive emotions, then, may offer particular benefits for patients with chronic pain who experience anhedonia[30], present with affective imbalance[36], or experience sleep disturbance[19–21], all of which are characterized by deficits in positive emotional function. These possibilities will of course need to be evaluated in future studies of Savoring.

Positive psychology interventions commonly employ a variety of skills (e.g., positive activity scheduling; gratitude journaling; best possible self imagery), and are usually deployed over the course of 6 or more sessions[35]. However, most of these interventions have not yielded superior effects to more general psychotherapeutic interventions for pain in trials to date[46]. One intervention, Mindfulness-Oriented Recovery Enhancement (MORE), has demonstrated reproducible, clinically significant effects on chronic pain outcomes that are mechanistically driven by increased positive emotionality and enhanced reward system function[25; 27; 29]. MORE employs a savoring skills practice that has been proposed to at least partially undergird the clinical effects observed[26]. However, because MORE also teaches other techniques, such as mindful breathing, positive reappraisal, and hypnosis, it has been difficult to isolate the specific therapeutic effects of savoring in trials to date. In contrast to broad-spectrum skill training approaches to pain psychotherapy, interventions that target a more limited set of therapeutic ingredients present an opportunity to evaluate therapeutic mechanisms more directly. Zeidan et al., for example, showed that brief mindful breathing training—without the introduction of other skills commonly taught in multicomponent mindfulness-based interventions—yields reproducible analgesic and mechanistic (e.g., orbital frontal cortex activation) effects that are distinguishable from an active, Slow Breathing control intervention[72]. By targeting a single skill and establishing clearly defined mechanistic outcomes, Savoring Meditation may be well-positioned to yield clinically meaningful pain-related benefits when delivered to patients suffering from deficits in the mechanisms targeted by this intervention approach.

In addition to boosting positive emotions and reducing anhedonia from pre-to post-intervention, Savoring produced substantial acute pain inhibition, with pain intensity ratings decreasing by 30% during Savoring compared to rest during the fMRI challenge. Slow Breathing was also analgesic, albeit to a lesser extent, yielding a 20% reduction in pain ratings. By comparison, a previous study with a similar noxious thermal stimulation paradigm showed that healthy subjects who employed mindful breathing evidenced 27% analgesia[72]. Thermal pain ratings following morphine administration tend to decrease between 5-10% relative to baseline[17; 54]. Coupled with the fact that no adverse events were reported in this study, it is evident that Savoring is a safe and effective strategy for both augmenting positive emotions and reducing acute experimental pain.

During the course of meditation and concomitant thermal stimulation, patients’ brains were scanned using arterial spin labeling to evaluate changes in CBF. We hypothesized that CBF in brain regions associated with the corticostriatal circuits would be altered by Savoring, and identified the NAc as the primary ROI because prior work showed that positive emotional music played during noxious thermal stimulation increased NAc BOLD activation in healthy participants [52]. However, the NAc was not significantly modulated by Savoring during noxious stimulation. The patients’ task in practicing Savoring was to use a memory of a personally meaningful experience that they associated with past positive emotions to generate a current positive emotional state, and then to maintain a present-moment awareness of that positive emotional state as a means of ‘regulating’ pain associated with a tonic noxious thermal stimulus. The null result for the NAc may have emerged because the NAc is perhaps most commonly activated during the *anticipation* of rewarding and aversive stimuli [10; 34; 42], and may be less involved in the integration of self-generated positive emotional experiences with pain perception. Although the NAc is clearly involved in both positive emotion and pain at a broad level, it may have been the wrong choice as the primary ROI for this specific experimental design.

Despite that null finding, whole brain analyses revealed significantly greater activation in a closely related corticostriatal region—the vmPFC—as well as increased functional connectivity between the vmPFC and the caudate while participants practiced Savoring during the administration of noxious thermal stimuli, relative to the practice of Slow Breathing. The observation of increased Savoring-induced CBF in the vmPFC during noxious stimulation comports with prior work that has converged on the vmPFC as both an important contributor to descending pain modulation[3; 33] and a neural hub of the generation of affective meaning out of self-referential value appraisals of contextual cues and memories of past experiences ([44; 45; 61], and for a review, see:[51]). Although the vmPFC is implicated in the integration and representation of both positive and negative emotional experiences, the observation that Savoring during noxious stimulation increased vmPFC connectivity with the caudate—a pivotal component of the striatum that assigns value to rewarding stimuli[32; 38]—suggests that Savoring recruits reward-enhancing corticostriatal circuits during painful experiences. This finding further supports prior work showing that positive autobiographical memories in healthy adults were associated with increased activation in both the mPFC and the caudate that corresponded to the magnitude of positive emotions experienced[57]. Relatedly, increases in corticostriatal[23] and electrocortical activity[27] during savoring of images representing natural rewards, has previously been linked with decreases in both chronic pain severity and anhedonia.

The present study had several limitations. First, the sample had relatively low clinical pain. Future studies will need to verify if the affective and neural responses to Savoring extend to patients with more severe chronic pain, and to clinical pain itself. Second, the sample was predominantly female, reflecting the general demographics of RA. Future studies will need to target chronic pain disorders with a more balanced sex/gender distribution. Third, because participants were instructed to avoid home practice during the duration of study participation, we were unable to evaluate practice adherence, or the effects of Savoring practice on pain and emotion in daily life. Future studies should use ecological momentary assessment to gather real-time data regarding meditation practice and its relationship to momentary changes in pain and pain-related measures. Fourth, although Savoring Meditation was designed as a “single skill” intervention that explicitly targets positive emotions, it utilized cognitive attention and mindfulness techniques to do so. Therefore we cannot rule out the possibility that there are additional “active ingredients” in Savoring Meditation—particularly cognitive effects—that contributed to the observed effects. Finally, because this was a pilot mechanistic trial, it was powered to detect large effects with a relatively small sample. Future studies will need to recruit a larger sample to evaluate the potential of Savoring to yield smaller, but still mechanistically and clinically meaningful effects.

### Conclusion

The present study showed that Savoring Meditation acutely decreased experimental pain sensitivity and engaged a vmPFC-caudate circuit during the application of noxious thermal stimuli in patients with generally well-controlled RA. Patients trained in Savoring Meditation also reported increased positive emotions and reduced anhedonic symptoms from pre-to post-intervention. Together, these findings suggest that Savoring Meditation may engage a self-referential, positive emotion-based descending modulatory mechanism through which patients self-manage pain. Future studies will need to test this hypothesis by extending the present findings to clinical outcomes in patients with more severe chronic pain.

## Data Availability

All data produced in the present study are available upon reasonable request to the authors, and execution of appropriate data sharing agreements with the institutions.

## Acknowledgments

This study was supported by a grant from the National Center for Complimentary and Integrative Health (NCCIH/NIH R61AT010134; PHF/DAS), as well as P30AR070254 (COB). The authors also wish to acknowledge the dedication and effort put forth by the patients, faculty, and staff of the Johns Hopkins Arthritis Center. Additionally, the authors wish to acknowledge the work of Kate Gilpin, MA, who assisted with organizing the medication data for this trial. The following potential conflicts of interest are acknowledged: PHF is on the scientific advisory board of Ninnion Therapeutics. DAS is an advisor to Empower Therapeutics. ELG is the Director of the Center on Mindfulness and Integrative Health Intervention Development. The Center provides Mindfulness-Oriented Recovery Enhancement (MORE), mindfulness-based therapy, and cognitive behavioral therapy in the context of research trials for no cost to research participants; however, Dr. Garland has received honoraria and payment for delivering seminars, lectures, and teaching engagements (related to training clinicians in MORE), including those sponsored by institutions of higher education, government agencies, academic teaching hospitals, and medical centers. Dr. Garland also receives royalties from the sale of books related to MORE. Dr. Garland has also been a consultant and licensor to BehaVR, LLC. All other authors declare no conflicts of interest.

## Notes

### Clinical Trial

NCT03975595

### Author Declarations

All participants provided informed consent, as required by the Johns Hopkins Institutional Review Board, which provided ethical approval for this study.

## References

[1] Anders B, Anders M, Kreuzer M, Zinn S, Fricker L, Maier C, Wolters M, Köhm M, Behrens F, Walter C. Sensory testing and topical capsaicin can characterize patients with rheumatoid arthritis. Clinical Rheumatology 2022;41(8):2351–2360.

[2] Arditte Hall KA, De Raedt R, Timpano KR, Joormann J. Positive memory enhancement training for individuals with major depressive disorder. Cognitive Behaviour Therapy 2018;47(2):155–168.

[3] Argaman Y, Kisler LB, Granovsky Y, Coghill RC, Sprecher E, Manor D, Weissman-Fogel I. The endogenous analgesia signature in the resting brain of healthy adults and migraineurs. The Journal of Pain 2020;21(7-8):905–918.

[4] Baliki MN, Geha PY, Fields HL, Apkarian AV. Predicting value of pain and analgesia: nucleus accumbens response to noxious stimuli changes in the presence of chronic pain. Neuron 2010;66(1):149–160.

[5] Barrett FS, Grimm KJ, Robins RW, Wildschut T, Sedikides C, Janata P. Music-evoked nostalgia: affect, memory, and personality. Emotion 2010;10(3):390.

[6] Bates D, Mächler M, Bolker B, Walker S. Fitting linear mixed-effects models using lme4. arXiv preprint arXiv:14065823 2014.

[7] Behzadi Y, Restom K, Liau J, Liu TT. A component based noise correction method (CompCor) for BOLD and perfusion based fMRI. Neuroimage 2007;37(1):90–101.

[8] Burns JW, Nielson WR, Jensen MP, Heapy A, Czlapinski R, Kerns RD. Specific and general therapeutic mechanisms in cognitive behavioral treatment of chronic pain. J Consult Clin Psychol 2015;83(1):1.

[9] Burns JW, Van Dyke BP, Newman AK, Morais CA, Thorn BE. Cognitive behavioral therapy (CBT) and pain education for people with chronic pain: Tests of treatment mechanisms. J Consult Clin Psychol 2020;88(11):1008.

[10] Carter RM, MacInnes JJ, Huettel SA, Adcock RA. Activation in the VTA and nucleus accumbens increases in anticipation of both gains and losses. Frontiers in behavioral neuroscience 2009:21.

[11] Cumming G. Understanding the new statistics: Effect sizes, confidence intervals, and meta-analysis: Routledge, 2013.

[12] Day MA, Jensen MP. Understanding pain treatment mechanisms: a new direction in outcomes research. Pain 2022;163(3):406–407.

[13] Day MA, Jensen MP, Ehde DM, Thorn BE. Toward a theoretical model for mindfulness-based pain management. The Journal of Pain 2014;15(7):691–703.

[14] Devilly GJ, Borkovec TD. Psychometric properties of the credibility/expectancy questionnaire. J Behav Ther Exp Psychiatry 2000;31(2):73–86.

[15] Edwards RR, Wasan AD, Bingham CO, Bathon J, Haythornthwaite JA, Smith MT, Page GG. Enhanced reactivity to pain in patients with rheumatoid arthritis. Arthritis research & therapy 2009;11(3):1–9.

[16] Ehde DM, Dillworth TM, Turner JA. Cognitive-behavioral therapy for individuals with chronic pain: efficacy, innovations, and directions for research. American Psychologist 2014;69(2):153.

[17] Fillingim RB, Ness TJ, Glover TL, Campbell CM, Hastie BA, Price DD, Staud R. Morphine responses and experimental pain: sex differences in side effects and cardiovascular responses but not analgesia. The Journal of Pain 2005;6(2):116–124.

[18] Finan PH, Garland EL. The role of positive affect in pain and its treatment. Clin J Pain 2015;31(2):177–187.

[19] Finan PH, Quartana PJ, Remeniuk B, Garland EL, Rhudy JL, Hand M, Irwin MR, Smith MT. Partial sleep deprivation attenuates the positive affective system: Effects across multiple measurement modalities. Sleep 2017;40(1):zsw017.

[20] Finan PH, Quartana PJ, Smith MT. The effects of sleep continuity disruption on positive mood and sleep architecture in healthy adults. Sleep 2015;38(11):1735–1742.

[21] Finan PH, Whitton AE, Letzen JE, Remeniuk B, Robinson ML, Irwin MR, Pizzagalli DA, Smith MT. Experimental sleep disruption and reward learning: moderating role of positive affect responses. Sleep 2019;42(5):zsz026.

[22] Fredrickson BL, Boulton AJ, Firestine AM, Van Cappellen P, Algoe SB, Brantley MM, Kim SL, Brantley J, Salzberg S. Positive emotion correlates of meditation practice: A comparison of mindfulness meditation and loving-kindness meditation. Mindfulness 2017;8:1623–1633.

[23] Froeliger B, Mathew A, McConnell P, Eichberg C, Saladin M, Carpenter M, Garland E. Restructuring Reward Mechanisms in Nicotine Addiction: A Pilot fMRI Study of Mindfulness-Oriented Recovery Enhancement for Cigarette Smokers. Evidence-Based Complementary and Alternative Medicine 2017;2017.

[24] Garland EL. Mindfulness-oriented recovery enhancement for addiction, stress, and pain: NASW Press, National Association of Social Workers, 2013.

[25] Garland EL, Atchley RM, Hanley AW, Zubieta J-K, Froeliger B. Mindfulness-Oriented Recovery Enhancement remediates hedonic dysregulation in opioid users: Neural and affective evidence of target engagement. Science advances 2019;5(10):eaax1569.

[26] Garland EL, Farb NA, Goldin PR, Fredrickson BL. The mindfulness-to-meaning theory: Extensions, applications, and challenges at the attention–appraisal–emotion interface. Psychological Inquiry 2015;26(4):377–387.

[27] Garland EL, Fix ST, Hudak JP, Bernat EM, Nakamura Y, Hanley AW, Donaldson GW, Marchand WR, Froeliger B. Mindfulness-Oriented Recovery Enhancement remediates anhedonia in chronic opioid use by enhancing neurophysiological responses during savoring of natural rewards. Psychol Med 2021:1–10.

[28] Garland EL, Hanley AW, Nakamura Y, Barrett JW, Baker AK, Reese SE, Riquino MR, Froeliger B, Donaldson GW. Mindfulness-oriented recovery enhancement vs supportive group therapy for co-occurring opioid misuse and chronic pain in primary care: A randomized clinical trial. JAMA Intern Med 2022;182(4):407–417.

[29] Garland EL, Hanley AW, Riquino MR, Reese SE, Baker AK, Salas K, Yack BP, Bedford CE, Bryan MA, Atchley R. Mindfulness-oriented recovery enhancement reduces opioid misuse risk via analgesic and positive psychological mechanisms: A randomized controlled trial. J Consult Clin Psychol 2019;87(10):927.

[30] Garland EL, Trøstheim M, Eikemo M, Ernst G, Leknes S. Anhedonia in chronic pain and prescription opioid misuse. Psychol Med 2020;50(12):1977–1988.

[31] Geschwind N, Peeters F, Drukker M, van Os J, Wichers M. Mindfulness training increases momentary positive emotions and reward experience in adults vulnerable to depression: a randomized controlled trial. J Consult Clin Psychol 2011;79(5):618.

[32] Grahn JA, Parkinson JA, Owen AM. The cognitive functions of the caudate nucleus. Progress in neurobiology 2008;86(3):141–155.

[33] Hadjipavlou G, Dunckley P, Behrens TE, Tracey I. Determining anatomical connectivities between cortical and brainstem pain processing regions in humans: a diffusion tensor imaging study in healthy controls. Pain 2006;123(1-2):169–178.

[34] Harris HN, Peng YB. Evidence and explanation for the involvement of the nucleus accumbens in pain processing. Neural regeneration research 2020;15(4):597.

[35] Hassett AL, Finan PH. The Role of Resilience in the Clinical Management of Chronic Pain. Current pain and headache reports 2016;20(6):1–9.

[36] Hassett AL, Simonelli LE, Radvanski DC, Buyske S, Savage SV, Sigal LH. The relationship between affect balance style and clinical outcomes in fibromyalgia. Arthritis Care & Research: Official Journal of the American College of Rheumatology 2008;59(6):833–840.

[37] Hausmann LR, Youk A, Kwoh CK, Gallagher RM, Weiner DK, Vina ER, Obrosky DS, Mauro GT, McInnes S, Ibrahim SA. Effect of a positive psychological intervention on pain and functional difficulty among adults with osteoarthritis: a randomized clinical trial. JAMA Netw Open 2018;1(5):e182533–e182533.

[38] Hikosaka O, Kim HF, Yasuda M, Yamamoto S. Basal ganglia circuits for reward value–guided behavior. Annual review of neuroscience 2014;37:289–306.

[39] Ibinson JW, Gillman AG, Schmidthorst V, Li C, Napadow V, Loggia ML, Wasan AD. Comparison of test–retest reliability of BOLD and pCASL fMRI in a two-center study. BMC Medical Imaging 2022;22(1):1–12.

[40] Jimenez SS, Niles BL, Park CL. A mindfulness model of affect regulation and depressive symptoms: Positive emotions, mood regulation expectancies, and self-acceptance as regulatory mechanisms. Personality and individual differences 2010;49(6):645–650.

[41] Kim HJ, Yang GS, Greenspan JD, Downton KD, Griffith KA, Renn CL, Johantgen M, Dorsey SG. Racial and ethnic differences in experimental pain sensitivity: systematic review and meta-analysis. pain 2017;158(2):194–211.

[42] Knutson B, Adams CM, Fong GW, Hommer D. Anticipation of increasing monetary reward selectively recruits nucleus accumbens. The Journal of neuroscience 2001;21(16):RC159.

[43] Kuznetsova A, Brockhoff PB, Christensen RH. lmerTest package: tests in linear mixed effects models. Journal of statistical software 2017;82:1–26.

[44] Lin W-J, Horner AJ, Burgess N. Ventromedial prefrontal cortex, adding value to autobiographical memories. Scientific reports 2016;6(1):1–10.

[45] Moore III WE, Merchant JS, Kahn LE, Pfeifer JH. ‘Like me?’: ventromedial prefrontal cortex is sensitive to both personal relevance and self-similarity during social comparisons. Social Cognitive and Affective Neuroscience 2014;9(4):421–426.

[46] Ong AD, Thoemmes F, Ratner K, Ghezzi-Kopel K, Reid MC. Positive affect and chronic pain: a preregistered systematic review and meta-analysis. Pain 2020;161(6):1140.

[47] Panagioti M, Gooding P, Tarrier N. An empirical investigation of the effectiveness of the broad-minded affective coping procedure (BMAC) to boost mood among individuals with posttraumatic stress disorder (PTSD). Behaviour research and therapy 2012;50(10):589–595.

[48] Parisi A, Roberts RL, Hanley AW, Garland EL. Mindfulness-oriented recovery enhancement for addictive behavior, psychiatric distress, and chronic pain: A multilevel meta-analysis of randomized controlled trials. Mindfulness 2022;13(10):2396–2412.

[49] Restom K, Behzadi Y, Liu TT. Physiological noise reduction for arterial spin labeling functional MRI. Neuroimage 2006;31(3):1104–1115.

[50] Riolo SA, Nguyen TA, Greden JF, King CA. Prevalence of depression by race/ethnicity: findings from the National Health and Nutrition Examination Survey III. Am J Public Health 2005;95(6):998–1000.

[51] Roy M, Shohamy D, Wager TD. Ventromedial prefrontal-subcortical systems and the generation of affective meaning. Trends in cognitive sciences 2012;16(3):147–156.

[52] Seminowicz DA, Remeniuk B, Krimmel S, Smith MT, Barrett FS, Wulff A, Furman AJ, Geuter S, Lindquist MA, Irwin MR. Pain-Related Nucleus Accumbens Function: Modulation by Reward and Sleep Disruption. PAIN 2019.

[53] Shin DD, Liu TT, Wong EC, Shankaranarayanan A, Jung Y. Pseudocontinuous arterial spin labeling with optimized tagging efficiency. Magnetic resonance in medicine 2012;68(4):1135–1144.

[54] Sibille KT, Kindler LL, Glover TL, Gonzalez RD, Staud R, Riley III JL, Fillingim RB. Individual differences in morphine and butorphanol analgesia: a laboratory pain study. Pain Medicine 2011;12(7):1076–1085.

[55] Singer JD, Willett JB. Applied longitudinal data analysis: Modeling change and event occurrence: Oxford university press, 2003.

[56] Snaith R, Hamilton M, Morley S, Humayan A, Hargreaves D, Trigwell P. A scale for the assessment of hedonic tone the Snaith–Hamilton Pleasure Scale. The British Journal of Psychiatry 1995;167(1):99–103.

[57] Speer ME, Bhanji JP, Delgado MR. Savoring the past: positive memories evoke value representations in the striatum. Neuron 2014;84(4):847–856.

[58] Speer ME, Delgado MR. Reminiscing about positive memories buffers acute stress responses. Nature Human Behaviour 2017;1(5):0093.

[59] Sturgeon JA, Finan PH, Zautra AJ. Affective disturbance in rheumatoid arthritis: psychological and disease-related pathways. Nature Reviews Rheumatology 2016;12(9):532–542.

[60] Suardi A, Sotgiu I, Costa T, Cauda F, Rusconi M. The neural correlates of happiness: A review of PET and fMRI studies using autobiographical recall methods. Cognitive, Affective, & Behavioral Neuroscience 2016;16:383–392.

[61] Summerfield JJ, Hassabis D, Maguire EA. Cortical midline involvement in autobiographical memory. Neuroimage 2009;44(3):1188–1200.

[62] Tabibnia G. An affective neuroscience model of boosting resilience in adults. Neuroscience & Biobehavioral Reviews 2020;115:321–350.

[63] Tan H, Maldjian JA, Pollock JM, Burdette JH, Yang LY, Deibler AR, Kraft RA. A fast, effective filtering method for improving clinical pulsed arterial spin labeling MRI. Journal of Magnetic Resonance Imaging: An Official Journal of the International Society for Magnetic Resonance in Medicine 2009;29(5):1134–1139.

[64] Trouvin A-P, Simunek A, Coste J, Medkour T, Carvès S, Bouhassira D, Perrot S. Mechanisms of chronic pain in inflammatory rheumatism: the role of descending modulation. Pain 2022:10.1097.

[65] Veehof M, Trompetter H, Bohlmeijer ET, Schreurs KMG. Acceptance-and mindfulness-based interventions for the treatment of chronic pain: a meta-analytic review. Cognitive behaviour therapy 2016;45(1):5–31.

[66] Wang Z. Improving cerebral blood flow quantification for arterial spin labeled perfusion MRI by removing residual motion artifacts and global signal fluctuations. Magnetic resonance imaging 2012;30(10):1409–1415.

[67] Wang Z, Aguirre GK, Rao H, Wang J, Fernández-Seara MA, Childress AR, Detre JA. Empirical optimization of ASL data analysis using an ASL data processing toolbox: ASLtbx. Magnetic resonance imaging 2008;26(2):261–269.

[68] Watson D, Clark LA. The PANAS-X: Manual for the positive and negative affect schedule-expanded form. 1999.

[69] Williams DR, Gonzalez HM, Neighbors H, Nesse R, Abelson JM, Sweetman J, Jackson JS. Prevalence and distribution of major depressive disorder in African Americans, Caribbean blacks, and non-Hispanic whites: results from the National Survey of American Life. Arch Gen Psychiatry 2007;64(3):305–315.

[70] Worsley KJ, Evans AC, Marrett S, Neelin P. A three-dimensional statistical analysis for CBF activation studies in human brain. Journal of Cerebral Blood Flow & Metabolism 1992;12(6):900–918.

[71] Yang Y. Social inequalities in happiness in the United States, 1972 to 2004: An age-period-cohort analysis. American sociological review 2008;73(2):204–226.

[72] Zeidan F, Emerson NM, Farris SR, Ray JN, Jung Y, McHaffie JG, Coghill RC. Mindfulness Meditation-Based Pain Relief Employs Different Neural Mechanisms Than Placebo and Sham Mindfulness Meditation-Induced Analgesia. J Neurosci 2015;35(46):15307–15325.

